# Sunlight exposure exerts immunomodulatory effects to reduce multiple sclerosis severity

**DOI:** 10.1101/2020.07.20.20157800

**Authors:** Patrick Ostkamp, Anke Salmen, Béatrice Pignolet, Dennis Görlich, Till F. M. Andlauer, Andreas Schulte-Mecklenbeck, Gabriel Gonzalez-Escamilla, Florence Bucciarelli, Isabelle Gennero, Johanna Breuer, Gisela Antony, Tilman Schneider-Hohendorf, Nadine Mykicki, Antonios Bayas, Florian Then Bergh, Stefan Bittner, Hans-Peter Hartung, Manuel A. Friese, Ralf A. Linker, Felix Luessi, Klaus Lehmann-Horn, Mark Mühlau, Friedemann Paul, Martin Stangel, Björn Tackenberg, Hayrettin Tumani, Clemens Warnke, Frank Weber, Brigitte Wildemann, Uwe K. Zettl, Ulf Ziemann, Bertram Müller-Myhsok, Tania Kümpfel, Luisa Klotz, Sven G. Meuth, Frauke Zipp, Bernhard Hemmer, Reinhard Hohlfeld, David Brassat, Ralf Gold, Catharina C. Gross, Carsten Lukas, Sergiu Groppa, Karin Loser, Heinz Wiendl, Nicholas Schwab, on behalf of the German Competence Network Multiple Sclerosis (KKNMS) and the BIONAT network

## Abstract

**Background:** Multiple sclerosis (MS) disease risk is associated with reduced sun exposure. This study assessed the relationship between measures of sun-exposure (vitamin D (vitD), latitude) and MS disease severity, the mechanisms of action, and effect-modification by medication and sun-sensitivity associated *MC1R* variants.

**Methods:** Two multi-center cohort studies (n_NationMS_=946, n_BIONAT_=991). Outcomes were the multiple sclerosis severity score (MSSS) and the number of Gd-enhancing lesion (GELs). RNAseq of four immune cell populations before and after UV-phototherapy of five MS patients.

**Results:** High serum vitD was associated with reduced MSSS (*P*_NationMS_=0.021; *P*_BIONAT_=0.007) and reduced risk for disease aggravation (*P*_NationMS_=0.032). Low latitude was associated with higher vitD, lower MSSS (*P*_NationMS_=0.018), fewer GELs (*P*_NationMS_=0.030) and reduced risk for aggravation (*P*_NationMS_=0.044). The influence of latitude on disability seemed to be lacking in the subgroup of interferon-β treated patients (interaction-*P*_BIONAT_=0.042, interaction-*P*_NationMS_=0.053). In genetic analyses, for carriers of *MC1R*:rs1805008(T), who reported increased sensitivity towards sunlight (*P*_NationMS_=0.038), the relationship between latitude und the number of GELs was inversed (*P*_NationMS_=0.001). Phototherapy induced a vitD and type I interferon signature that was most apparent in the transcriptome of monocytes (*P*=1×10^−6^).

**Conclusion:** VitD is associated with reduced MS severity and disease aggravation. This is likely driven by sun-exposure, as latitude also correlated with disability and serum vitD. However, sun-exposure might be detrimental for sun-sensitive patients. A direct induction of type I interferons through sun-exposure could explain a reduced effect of latitude in interferon-β treated patients. This could also explain opposite effects of sun-exposure in MS and the type I interferon and sun-sensitivity-associated disease Lupus.

## Introduction

Multiple sclerosis (MS), characterized by demyelinating lesions, is the most common neuroinflammatory disease of the central nervous system and presumably of autoimmune origin (1). In most cases, the disease is diagnosed at a young age, predominantly occurs in women, and follows a relapsing-remitting course, which can be superseded by a secondary, progressive stage (2). Treatment options include interferon-β (IFN-β) (a type I interferon normally involved in the defense against viral infections), monoclonal antibodies against leukocyte migration-mediating adhesion molecules, and other immunomodulatory drugs (3). Etiologically, environmental factors have been shown to play an important role (4) and insufficient sunlight exposure has been suspected to be critical for the *initial development* of MS (5). The best characterized mediator of ultraviolet radiation (UVR)-dependent effects is vitamin D (vitD), that is generated from its precursor 7-dehydrocholesterole in the skin, further metabolized in the liver and in the kidney, and that exerts its function in its active form 1-α,25-dihydroxyvitamin D_3_ (1α,25(OH)_2_D_3_), also known as calcitriol (6). Precursors of active vitD can also be found in food in the form of ergocalciferol (or vitamin D_2_), which is however of little relevance for total vitD serum levels (7). For MS, low vitD levels have been shown to be associated with disease risk (8) and mendelian randomization studies hint towards a causal role for vitD (9, 10). However, it is possible that alternative ultraviolet radiation dependent pathways play a role, as well (11). Furthermore, it is still a topic of debate whether UVR-/vitD only modulate *disease risk*, or if *disease severity* is affected, as well. In mouse models, UVR was shown to ameliorate disease severity (12). Human observational studies suggested an influence of vitD on disease activity (13, 14) and prospective trials also suggested an effect on the formation of lesions, but the primary endpoints of supplementation studies have not been met so far, raising doubt about the role of vitD (15-17). It has also been argued that reverse causation could influence the results from observational studies, i. e. low vitamin D could be caused by disease activity rather than *vice versa* (5). Reports on the effect of latitude (which can be regarded as an independent measure of sun-exposure) on disease severity are scarce and could either not identify any effects or showed contradictory effects (18, 19). Recently, studies also suggested vitD-independent effects of UVR in MS (20, 21). Besides the assumed main effects of UVR/vitD on MS severity, there might be factors that could alter the effects of UVR-/vitD. Medication -especially IFN-β therapy - has been suggested to modulate vitD-production, and the correlation between vitD and disease activity has been shown to disappear after IFN-β treatment-onset (22, 23). Another potentially modifying factor of UVR-effects is the melanocortin 1 receptor (MC1R). The MC1R is responsible for melanin synthesis upon UVR exposure and carrying loss of function variants results in a red hair and fair skin phenotype with increased sun-sensitivity (24, 25). The MC1R is also known to confer immunosuppressive effects and functional signaling ameliorates disease course in mouse models of MS (26). Moreover, the MC1R agonist adrenocorticotropic hormone (ACTH) is used to treat disease exacerbations in MS and part of the effect of ACTH could be mediated through MC1R (27). Therefore, the MC1R could be modifier of UVR-effects in MS.

This study aims to provide a better understanding of the effects of UVR/vitD on MS severity, to dissect the mechanistic network involved in this, and to deduce the role of modulatory factors like medication & sun-sensitivity associated genotypes. In the setting of two large, independent multicenter studies the effects of the sun-exposure measures vitD and latitude on disease severity were investigated. To assess the role of further mechanistic pathways in mediating-/modifying UVR or vitD effects, interactions between sun-exposure measures and either medication or *MC1R* genotype were assessed. Finally, in an unbiased approach the UVB-induced transcriptomic changes in immune cells of MS patients from our 2014 pilot study were assessed, illuminating the pathways triggered by UVR.

## Methods

### Patients

Data from the *NationMS* cohort, a prospective multicenter observational study, were used. In total, clinical data of 946 clinically isolated syndrome (CIS) / relapsing remitting multiple sclerosis patients (RRMS) recruited between 2010-2017 in 21 centers in Germany, were obtained (28). Patients were followed up and longitudinal data is available for 798 patients (Fig. 1). Included were patients with age ≥ 18, who were diagnosed with either clinically isolated syndrome with fulfillment of three Barkhof criteria or confirmed relapsing remitting MS according to revised 2005 McDonald criteria. All patients were treatment-naïve at baseline with onset of symptoms no longer than 2 years before inclusion. The study was approved by the lead ethics committee (Ethik-Kommission der Ruhr-Universität Bochum, registration number: 3714-10) and all local committees. Informed written consent was obtained from all participants and the study was performed according to the Declaration of Helsinki. For replication, data of 990 patients from the *BIONAT* cohort (Study identifier: NCT00942214), a French, multicenter, cohort, at their baseline assessment, were acquired (29). *BIONAT* patients were not treatment-naïve before inclusion. Study investigators for *NationMS* and *BIONAT* can be found in the supplementary material. For RNA sequencing, peripheral blood mononuclear cell samples (PBMC) of five patients from our 2014 pilot study were used (12).

**Figure 1:**
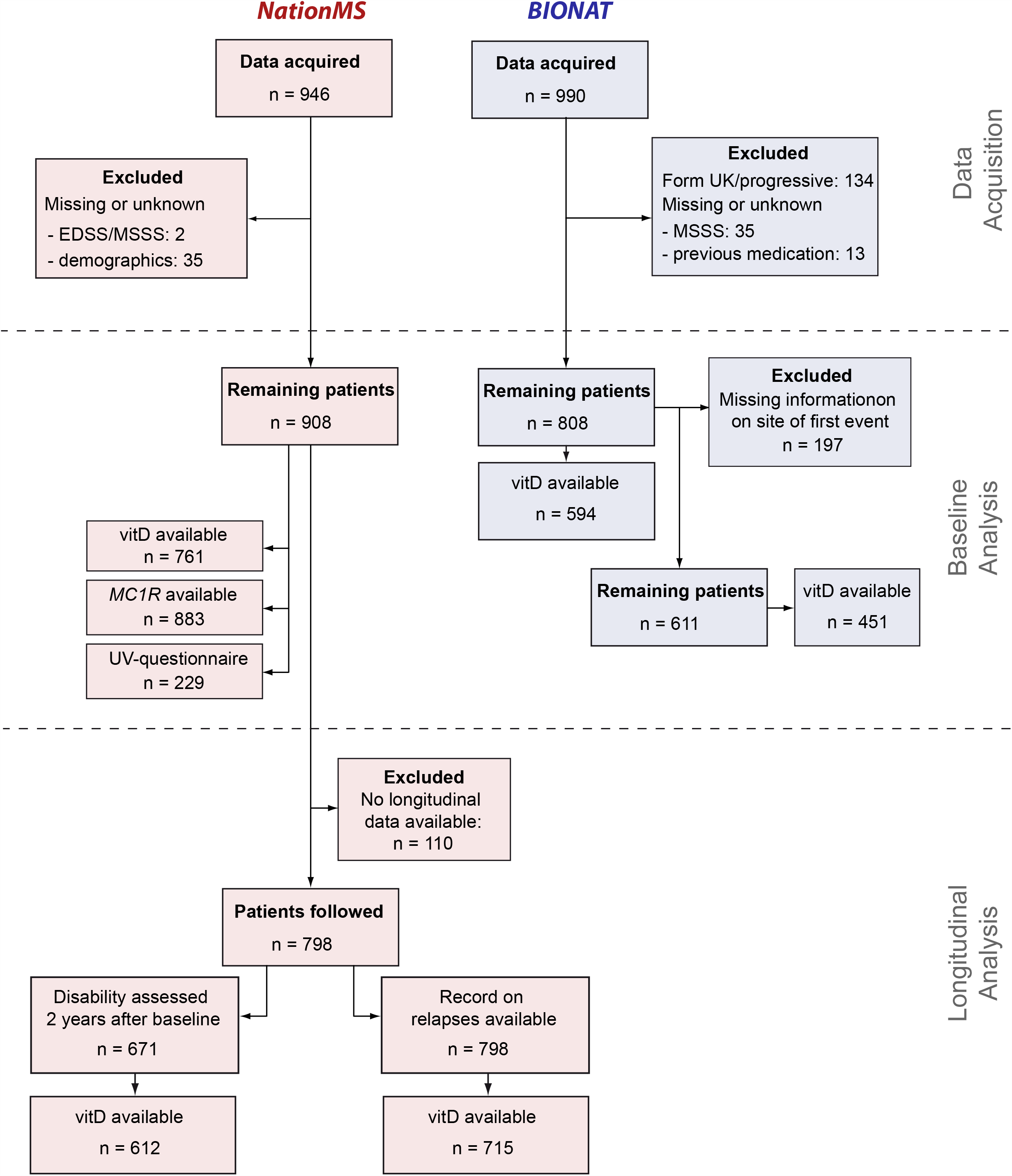
Study Flow-Chart. Data of 946 therapy-naïve patients were acquired for the *NationMS* cohort and data of 990 *BIONAT* patients with a history of previous medication for multiple sclerosis have been acquired. Datasets were filtered for missing information with 908 *NationMS* and 808 *BIONAT* patients remaining at baseline. For the analysis of disease severity *BIONAT* patients were further excluded when information on site of first event was unknown/unavailable (remaining total n=611, n for remaining patients with available information on serum vitamin D = 451). To assess the influence of latitude on serum vitD levels this information was not necessary and n=594 patients were assessed. For *NationMS*, longitudinal information on relapses was available for n=798 and information on disability two years after baseline was available for 671 patients.

### Genotyping and estimation of population stratification

Genotyping and quality control was performed as previously published (30, 31). Briefly, genotyping was performed using an Illumina OmniExpress chip and quality control (QC) was performed in PLINK v1.90. Genotypes for the most common *MC1R* variants were extracted (rs1805008:C>T, rs885479:G>A, rs2228479:G>A), and population stratification was calculated by multidimensional scaling (MDS) as previously published (30).

### Serum vitamin D measurements

For *NationMS*, vitamin D was measured as the serum level of 25(OH)D_3_. Measurement was performed in *NationMS* for n=761 patients with a commercially available kit for liquid chromatography-mass spectrometry (LC-MS/MS) (Recipe, Munich, Germany) on a Shimadzu 8040 LC–MS/MS instrument coupled to a UHPLC system (Nexera, Shimadzu). For *BIONAT*, total 25(OH)D_2+3_measurements were available for 579 patients. Total vitD has been measured using a commercially available electro-chemiluminescence competitive 25-hydroxyvitamin D binding assay (Cobas, Roche), and quantified on a Roche Cobas 8000 e602 analyzer (Roche Diagnostics, Mannheim, Germany). As food-derived vitD_2_ only contributes little to total serum vitD levels, there are only minimal differences between total vitD and vitD_3_ serum levels.

### Questionnaire on skin type

Two hundred twenty-nine patients of the *NationMS* cohort were asked to report their skins potential reaction to sunlight at noon for one hour without the use of sunscreen in summer. Possible answers were ‘Tan’, ‘Sunburn with a delayed tan’, ‘Sunburn without tan and without blisters’ and ‘Sunburn without tan and with blisters’. Patients were asked to report skin reddening, a feeling of heat and itching after 12-24 hours as a sunburn, and liquid-filled skin alterations with a diameter ≥5 mm as blisters. The resulting variable was treated as ordinal.

### Disease severity outcomes

The multiple sclerosis severity score (MSSS) according to Roxburgh (32) was used as the primary measure of disability in both cohorts. Additionally, for the *NationMS* cohort, magnetic resonance imaging (MRI) was carried out according to standardized protocols (33) and assessed for gadolinium (Gd)-enhancing lesions in T1-weighted images. Electromagnetic Field strength was 3 Tesla across all centers and evaluation was carried out at each site separately by certified neuro-radiologists. To assess disability accumulation over time in the *NationMS* cohort, the change in the expanded disability status scale (EDSS) was assessed between follow up 2 (T2, 2 years from baseline) and baseline visit. Furthermore, relapses during the course of the study were recorded.

### Statistical analysis

The MSSS was modelled as a continuous variable and the influence of the UVR-exposure-measures, latitude and vitD levels, was assessed using linear models. In the *NationMS* cohort these analyses were adjusted for age, sex, body mass index (BMI, weight/height^2^), smoking (yes/no), alcohol (yes/no), month of assessment (treated as a categorical variable with 12 factor-levels), clinical subtype (CIS or RRMS) and the site/symptom of first disease manifestation (categorical). A random intercept was used to adjust for center variability, which was omitted when assessing the influence of latitude (latitude is intrinsic to the variability between centers and inclusion would lead to unresolvable multi-collinearity). For the *BIONAT* cohort, these analyses were adjusted for age, sex, month of assessment, site/symptom of first manifestation, and prior medication (other covariates not available for *BIONAT*). Center variability was treated as in the *NationMS* cohort. For *BIONAT*, also the interaction of vitD /-or latitude and prior treatment was tested, followed by subgroup analyses to obtain subgroup specific estimates. The number of Gd-enhancing lesions in *NationMS* was modelled using a zero-inflated negative binomial-model to account for overdispersion. Confounder-adjustment was done as in the linear model. Plots for Gd-enhancing lesions include a left y-axis, showing the observed number of lesions, and a right y-axis, showing the mean number of lesions over x. The influence of the *MC1R* genotype on skin reaction to sun exposure was modelled using cumulative link models (CLMs, ‘ordinal regression’) and adjusted for age, sex, BMI, alcohol consumption, smoking, population stratification (first three MDS components). To assess whether *MC1R* genotype modifies the effect of UVR exposure on severity, an interaction-term between *MC1R* genotype and measures of sun-exposure (latitude or vitD) was added to the models for disease severity and adjusted as the prior models for disease severity plus population stratification. To assess the influence of vitD and latitude on the risk for relapses in the *NationMS* cohort, (mixed effects-) cox-regression was performed and adjusted as the baseline models for disease severity plus medication after baseline (therapy was initiated after baseline visit). The difference between the EDSS two years after baseline and the baseline EDSS was calculated and declared as ΔEDSS. The ΔEDSS was modelled using linear and linear mixed models and adjusted as the baseline model plus the type of medication initiated after baseline visit. Additionally, the models were adjusted for the number of lesions and the EDSS at baseline to account for differences in baseline severity. Model fit was assessed by inspection of scaled quantile residuals (34). Model coefficients, confidence intervals and *P*-values were reported from fully adjusted models with significance threshold of α = 0.05 using two-sided tests. Genetic analyses of the *MC1R* genotype that were not of confirmatory nature (i. e., assessing disease severity and not skin reaction), were assessed for genome-wide significance (α = 5×10^−8^).

### RNA sequencing analysis

RNA-sequencing (RNAseq) was performed using samples of five patients from our 2014 pilot study (35). Frozen PBMC samples from before and after six weeks of phototherapy (ten samples in total) were thawed and stained for CD3, CD4, CD8, CD14, CD19 and CD56 (all antibodies from Biolegend). Viable cells were identified by forward and sideward scatter characteristics. T-cell subsets were identified as CD56-CD3+ cells being either CD4+CD8- or CD8+CD4-cells. B-cells were identified as CD3-CD56-CD14-CD19+, and monocytes were identified as CD3-CD19-CD56-CD14+ cells. Cells were sorted on a FACS Aria III machine and subjected to RNA isolation. RNA sequencing was performed on an Illumina NextSeq 500. QC was done using *FastQC* and read quantification was done in *Kallisto (36)*. Differential gene expression analysis was performed using the quasi-likelihood approach in *edgeR (37)*. To test the enrichment of the vitD- and type I interferon-associated genes, distribution-free permutation tests were performed (38). Due to the limited sample size and the exploratory nature of this part of the study, genes with *P*-values < 0.05 were used for enrichment tests. Genes associated with the respective pathways were extracted from *wikipathways* (39), collapsing the quality approved gene-/protein lists from the pathways ‘Vitamin D receptor-pathway’ (WP2877) and ‘Non-genomic effects of vitamin D’ (WP4341) to generate a reference set of vitD-associated genes. For the type I interferon pathway, the gene-/protein lists from the ‘Type I interferon signaling pathway’ (WP585) and the ‘DDX58/IFIH1-mediated induction of interferon-alpha/beta’-pathway (WP1904) were collapsed to generate a reference set of type I interferon-associated genes. For permutation tests, the overlap of genes regulated by phototherapy (*P*<0.05) for each respective cell type with the reference gene-set was calculated. Next, random samples of the same size as the list of genes with *P*<0.05 were taken from the list of all genes expressed by the respective cell type, and the overlap with the reference gene-set was calculated. To obtain a *P*-value the random sampling was repeated 100,000 times and the number of occasions when randomly sampled genes overlapped equally or more than the test-set with the reference sets was divided by the number of permutations (n=100,000) (38).

### Software

Statistical analyses were conducted in *R* v3.6.0 using the packages *stats, lme4, glmmTMB, ordinal* and *DHARMa*. To extract and process data from NASA’s OMI dataset the package *RNetCDF* was used. The remaining data were manipulated and plotted using the *R* packages *tidyR, ggplot2* and *RColorBrewer*. For population stratification estimation, *PLINK* v1.90b5.2 was used. For analysis of RNAseq data, *Kallisto and edgeR* were used. For pathway extraction from wikipathways the package *rWikiPathways* was used.

## Data availability

Data and *R* code is available from the authors upon reasonable request. Full results from RNAseq can be found in the supplementary material.

## Results

### Cohort Characteristics

For the *NationMS* cohort, data of 946 treatment-naïve patients at baseline with CIS or RRMS were acquired (duration < 2 years). The study procedure is shown in (Fig. 1). Baseline characteristics are described in Table 1. Genotyping for *MC1R* was successful and passed QC for 883 (97.25%) patients of the 908 patients that had full demographic and clinical information available. The most common *MC1R* variant allele was rs1805008:T (n=148/883, 16.76% with ≥ 1 risk allele, minor allele frequency (MAF)=8.60%), followed by rs2228479:A (n=140/883, 15.86% with ≥ 1 risk allele, MAF=8.10%), and rs885479:A (n=76/883, 8.61% with ≥ 1 risk allele, MAF=4.42%). Patients were subjected to regular follow-ups. In total, 798 patients were longitudinally followed (Fig. 1). Of these, 155 (19.42%) patients remained untreated after baseline assessment while 643 (80.58%) received treatment. The most prescribed medication was IFN-β (n=359, 44.99 %) (Table 2). Moreover, 355 patients (44.49%) reported a relapse in the time after baseline assessment with a mean time to relapse of 360.34 (SD=359.55) days. For another 671 patients, disability was assessed two years after baseline, and 156 patients (22.38%) had an increase of at least one point on the EDSS.

**Table 1:** Cohort baseline characteristics

|  | ***NationMS*** | ***BIONAT*** |
| --- | --- | --- |
| **Number of patients** | 908 | 808 |
| **Median Age (IQR)** | 32.43 (26.76 to 41.15) | 37 (30 to 43) |
| **Median BMI** | 24.22 (21.56 to 27.64) | - |
| **Male (%)** | 271/908 (29.85) | 196/808 (24.26) |
| **Smokers (%)** | 305/908 (33.59) | - |
| **Consume alcohol (%)** | 674/908 (74.23) | - |
| **RRMS (%)** | 485/908 (53.42) | 808/808 (100) |
| **Mean 25(OH)D<sup>a</sup>** | 21.56 ng/mL | 20.95 ng/mL |
| **Median MSSS (IQR)** | 4.31 (2.83 to 5.58) | 5.23 (3.41 to 6.92) |
| **≥ 1 Gd-enhancing lesion (%)** | 350 (38.55) | - |
| **Most common site / symptom of first manifestation<sup>a</sup> (%)** | Sensory system (411/908, 45.26%) | Spinal cord (138/611, 22.59 %) |

Abbreviations: 25(OH)D = 25-hydroxy-vitaminD, BMI = body mass index (weight/height<sup>2</sup>), Gd = Gadolinium, IQR = interquartile range, MSSS = multiple sclerosis severity score, RRMS = relapsing-remitting multiple sclerosis. <sup>a</sup> According to the number of patients with available data (see Figure 1)

**Table 2:** Medication before-/after first assessment

| ***NationMS*** |  |  |
| --- | --- | --- |
| **Medication after first assessment** | **Number** | **Percent (of 798 total)** |
| Fingolimod | 21 | 2.63 |
| Fumarat | 64 | 8.02 |
| Glatiramer acetate | 162 | 20.30 |
| Interferon-β | 359 | 44.99 |
| Monoclonal antibodies<sup>a</sup> | 23 | 2.88 |
| Naïve | 155 | 19.42 |
| Teriflunomide | 14 | 1.75 |
| ***BIONAT*** |  |  |
| **Medication before first assessment** | **Number** | **Percent (of 808 total)** |
| Glatiramer acetate | 236 | 29.21 |
| Interferon-β | 483 | 59.78 |
| Naïve | 89 | 11.01 |

<sup>a</sup> The monoclonal antibodies Natalizumab, Rituximab and Alemtuzumab were grouped due to low frequencies

For the *BIONAT* cohort, data of 990 MS patients at their baseline assessment were acquired (Fig. 1). Full information on demographics, clinical subtype, medication and MSSS was available for 808 (81.62%). Most patients had already received therapy before the baseline assessment. The most prescribed medication was IFN-β (59.77%) (Table 2).

Characteristics of the five patients treated with UVB-phototherapy were previously published (12). Two of the patients were treated with IFN-β, one with GA, one with Natalizumab and one had not received any treatment.

### Low vitamin D levels and high latitude are associated with clinical disease severity

Based on the available evidence before start of the study, the association between latitude, vitD and clinical severity was assessed (Fig. 2). In the *NationMS* cohort vitD levels were associated with lower disability, as assessed by the MSSS (β=-0.014, 95% Confidence interval (CI)=[-0.026 to −0.002], *P*=0.021). Higher latitude (corresponding to lower sun exposure) was associated with worse MSSS scores in the *NationMS* cohort (β=0.092, 95% CI=[-0.016 to 0.168], *P*=0.018) (Fig 2 A). The risk for Gd-enhancing lesions increased by 8.31% for every one degree increase in latitude (RR=1.08, 95% CI=[1.01 to 1.16], *P*=0.030) (Fig. 2 B). Connecting these two results, lower latitude was expectedly associated with higher vitD levels (β=-0.56, 95% CI=[-1.030 to −0.090]) (Supplementary Fig. 1).

**Figure 2:**
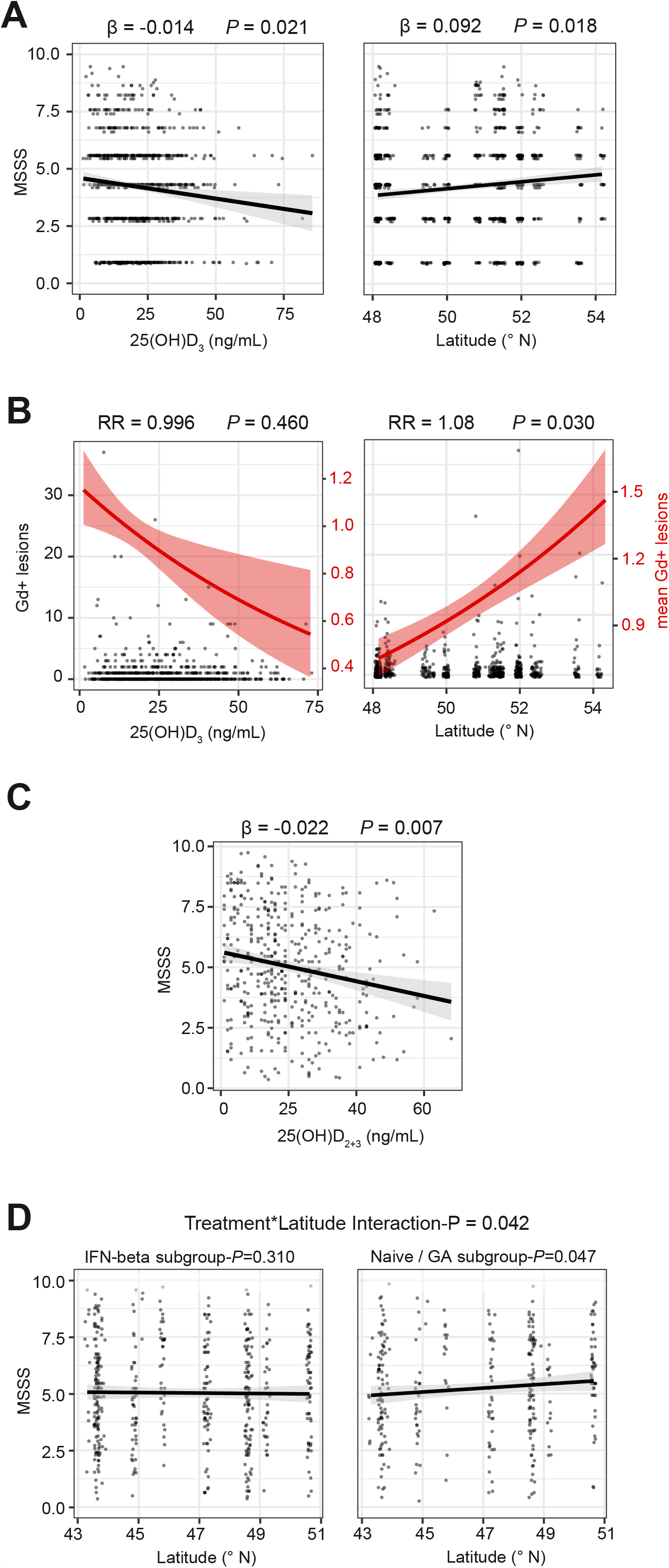
Influence of sun exposure measures on clinical severity. **A** Dotplots for multiple sclerosis severity score (MSSS) in relation to 25(OH)D_3_ levels and latitude (of the patients’ medical center) with least squares linear regression lines ± standard error for *NationMS* (n_vitD_=761, n _lat_=908). **B** Dotplot for gadolinium (Gd)-enhancing lesions in relation to 25(OH)D_3_ levels (n=761) and latitude (n=908) for the *NationMS* cohort. Left y-axis corresponds to the dotplot displaying observed counts, right y-axis corresponds to the red line displaying mean number of lesions ± standard error. **C** Dotplot for MSSS in relation to 25(OH)D_2+3_ with least squares linear regression line ± standard error for *BIONAT*. **D** Dotplots for MSSS in relation to latitude stratified by previous treatment (n_IFN-β_=363, n_Naive/GA_=248) with least-squares regression line ± standard error. Analyses for *NationMS* are adjusted for age, sex, body mass index, smoking, alcohol consumption, clinical subtype, neurological site of first manifestation, month of assessment and center. Analyses for *BIONAT* are adjusted for age, sex, neurological site of first manifestation, month of assessment and center. Adjustment for center was omitted when analyzing the effect of latitude.

### The effect of latitude is absent in patients treated with Interferon-β

The hypotheses were then also tested in the *BIONAT* cohort. Here, higher vitD levels showed an association with lower disability, as well (β=-0.022, 95% CI=[-0.037 to −0.007], *P*=0.007) (Fig. 2 C). However, in contrast to the results from the *NationMS* cohort, there was no clear trend for an effect of latitude on the MSSS (β=-0.004, 95% CI=[-0.076 to 0.068], *P*=0.913). As not all *BIONAT* patients were therapy-naïve at baseline and IFN-β therapy has been shown to cloud the effects of vitD, it was hypothesized that the missing effect of latitude could also be due to confounding by IFN-β therapy (40). Interaction-analyses indeed showed that the effect of latitude on disability was significantly different depending on IFN-β treatment status (β=0.145, 95% CI[0.005 to 0.283], interaction-*P*=0.042), and subsequent subgroup analyses confirmed that higher latitude was associated with increased MSSS in patients whose last medication was not IFN-β (β=0.115, 95% CI[0.001 to 0.227], *P*=0.047), while patients treated with IFN-β did not show this effect (β=-0.048, 95% CI[-0.141 to 0.045], *P*=0.310) (Fig. 2 D). Consistent with the results from the *NationMS* cohort, also in the *BIONAT* cohort lower latitude was associated with higher vitD levels (β=-0.44, 95% CI=[-0.87 to −0.018], *P*=0.041), and in line with previous reports IFN-β-treated patients had higher vitD levels than therapy-naïve patients (β=4.020, 95% CI[0.450 to 7.560], *P*=0.031) (Supplementary Fig. 1) (22).

### High Vitamin D and low latitude are associated with reduced risk for relapses and disability accumulation

To further test the predictive capacity of vitD and latitude with regard to disease burden accumulation, data on confirmed relapses after baseline in the *NationMS* cohort were assessed (Fig. 3). In a mixed effects cox regression vitD (treated as continuous variable) reduced the risk for a relapse by 1% for every 1 ng/mL of serum vitD (hazard ratio (HR)=0.99, 95% CI=[0.978 to 0.999], *P*=0.044). For the purpose of visualization vitD was grouped into three categories: 1) the 20% of patients with the highest vitD levels (≥30.31 ng/mL); 2) the 20% of patients with the lowest vitD levels (≥10.86 ng/mL); and 3) the patients in-between (Fig. 3 A). This categorization is also close to common definitions of optimal (≥30 ng/mL) and deficient (<10 ng/mL) vitD levels (41, 42). Furthermore, baseline vitD was inversely associated with a lower ΔEDSS (β=-0.010, 95% CI=[-0.019 to −0.001], *P*=0.031) (Fig. 3 B). Latitude showed no significant effect on the risk for relapses in an unstratified analysis (Fig. 3 C). If stratified by medication as in the *BIONAT* cohort, patients at lower latitudes trended towards a lower risk for relapses when they were untreated or received GA (HR=0.937, 95% CI[0.835-1.042], *P*=0.22), while there was no clear trend for IFN-β treated patients (HR=0.964, 95% CI[0.881-1.055], *P*=0.427) (Fig. 3 D+E). Regarding changes in disability, lower latitude was associated with a lower ΔEDSS (β=0.67, 95% CI=[0.013 to 0.124], *P*=0.017) and if stratified by medication, the effect seemed to be stronger in patients who were not treated with IFN-β (Naïve-/GA-subgroup: β=0.124, 95% CI=[0.004 to 0.244], *P*=0.043; IFN-β-subgroup: β=0.024, 95% CI=[-0.058 to 0.107], *P*=0.556), although this was of borderline-significance (interaction-*P*=0.053) (Fig. 3 F-H).

**Figure 3:**
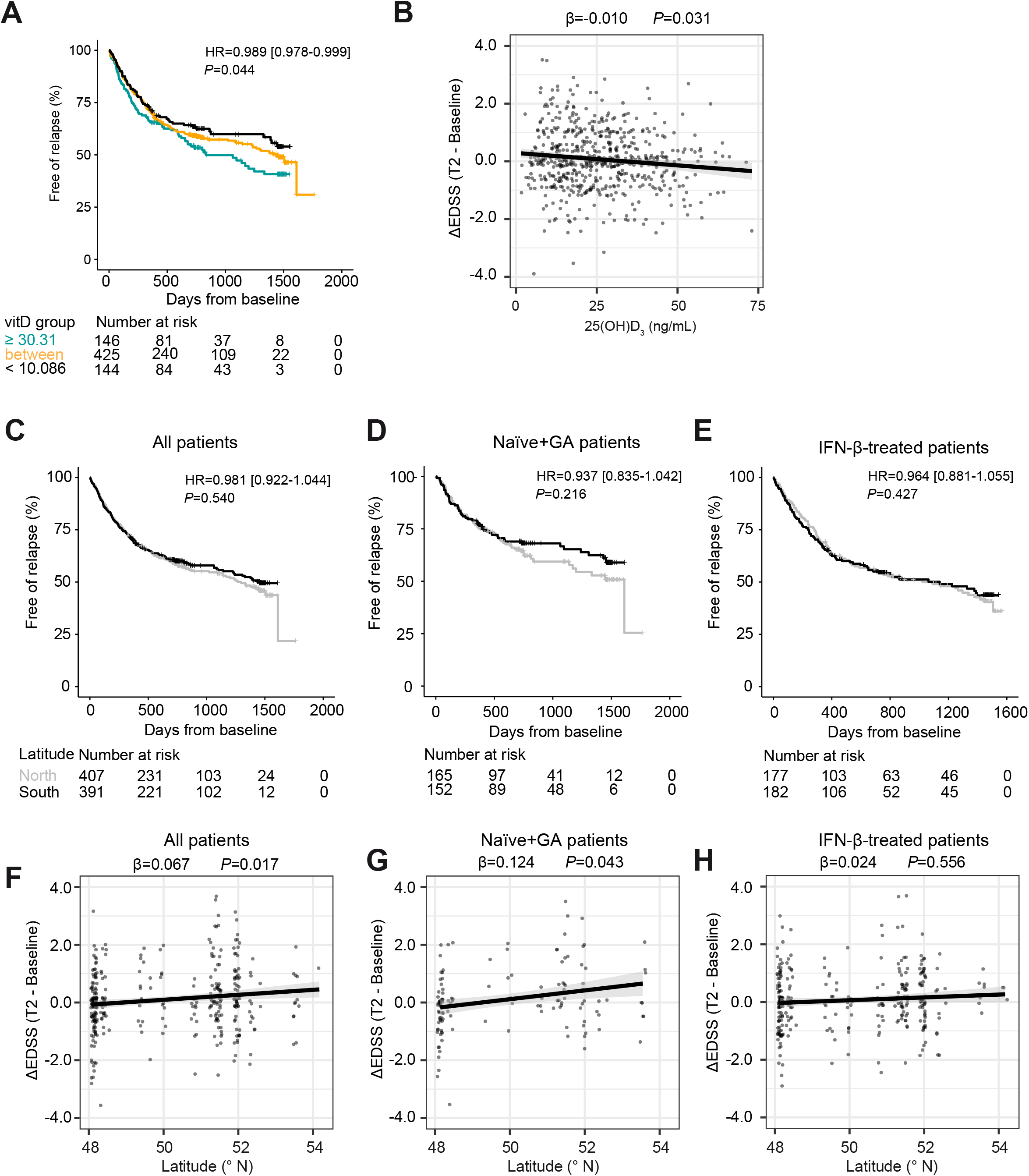
Influence of vitamin D and latitude on risk for relapses and disability accumulation. **A** Time-to-event curves displaying the proportion of relapse-free patients over time for the *NationMS* cohort grouped by serum 25(OH)D_3_ levels (Colorcode: Jade=The 20% of patients with the lowest 25(OH)D_3_ levels, cutpoint: <10.086 ng/mL; Black=the 20% of patients with the highest 25(OH)D_3_ levels, cutpoint: ≥30.31 ng/mL; Orange=patients in between) and complemented with a table displaying the number at risk over time. **B** Dotplot for the change in EDSS (ΔEDSS) in relation to baseline 25(OH)D_3_ levels with least squares linear regression line ± standard error. **C-D** Time-to-event curves displaying the proportion of relapse-free patients over time for the *NationMS* cohort grouped by latitude (north defined as ≥ median latitude within the cohort = 50.85 °N) and complemented with a table displaying the number at risk over time. D and E display the results for the analyses if stratified by medication. **F-H** Dotplot for the ΔEDSS in relation to latitude with least squares regression line ± standard error. G and H display the results for the analyses if stratified by medication. Analyses are adjusted for age, sex, body mass index, smoking, alcohol consumption, clinical subtype, neurological site of first manifestation, month of assessment, medication after baseline assessment and center. Differences in baseline-severity were adjusted by using the baseline MSSS and the number of lesions at baseline as covariates. Adjustment for center was omitted when analyzing the effect of latitude.

### Sun-sensitivity-associated MC1R missense variants might modify UVR-mediated effects

*MC1R* missense variants strongly increase an individual’s sensitivity to UVR and could, therefore, modify the beneficial effect of UVR on MS disease severity. First, the previously described influence of *MC1R* genotype on sun sensitivity was confirmed using ordinal regression (Fig. 4 A-C). Whereas there was no statistically significant effect for the low-penetrance variants rs885479 (OR=2.05, 95% CI=[0.764 to 5.541], *P*=0.153) and rs2228479 (OR=0.97, 95% CI=[0.492 to 1.911], *P*=0.936), the T allele of the high-penetrance variant rs1805008 increased the odds for a severe reaction to sunlight by 91.5% (OR=1.92, 95% CI=[1.040 to 3.560], *P*=0.038) (Fig. 4 A). Next, as rs1805008:T carriers reported severe reactions to sunlight, the effect of an interaction-term between measures of sun exposure (latitude, vitD) and rs1805008 on clinical severity was assessed (at baseline). No significant interactions were found regarding the MSSS (Fig. 4 D). The interaction term of rs1805008:T and latitude was nominally significant in the analysis of Gd-enhancing lesions (interaction-*P*=0.001). In subgroup analyses for carriers of rs1805008:T, the risk for Gd-enhancing lesions increased by 20.5% for every 1° decrease in latitude (RR=1.21, 95% CI=[1.010 to 1.437], *P*=0.034), while it was reduced by 11.6% in non-carriers for every 1° decrease in latitude (RR=0.88, 95% CI=[0.808 to 0.947], *P*=9.2×10^−4^) (Fig. 4 E). Both subgroup analyses were nominally significant. Inconsistencies between read-outs might be due missing correlation between Gd-enhancing lesions and the MSSS (ρ=0.03, *P*=0.33). No interaction between rs1805008:T and vitD levels regarding Gd-enhancing lesions was found, though (RR=1.00, 95% CI=[0.968 to 1.137], *P*=0.825).

**Figure 4:**
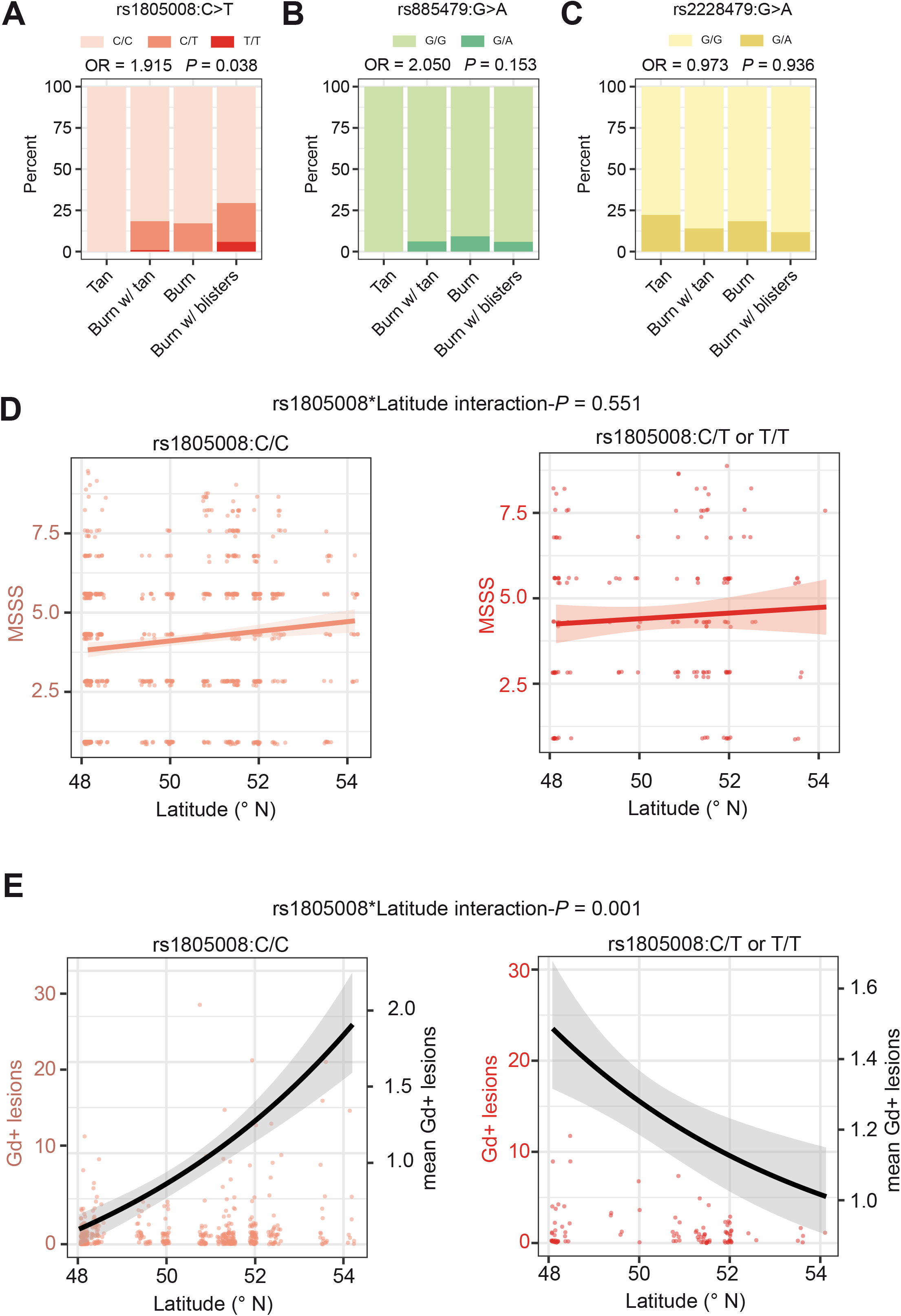
Interaction of *MC1R* genotype and sun exposure. **A-C** Barplots displaying the fraction of total patients (n_total_=229) who reported their reactions to sun exposure at noon in summer for the *MC1R* genotypes **A** rs1805008(C>T), **B** rs885479(G>A) and **C** rs2228479(G>A). This analysis was adjusted for age, sex, body mass index, smoking, alcohol consumption and population stratification. **D** Dotplots for MSSS in relation to latitude with leastsquares linear regression line stratified by rs1805008 genotype. **E** Dotplots for gadolinium-enhancing lesions in relation to latitude (left y-axis) complemented with the mean number of lesions (black line, right y-axis). Analyses for **D & E** were adjusted for age, sex, body mass index, smoking, alcohol consumption, clinical subtype, neurological site of first manifestation, month of assessment and center. Adjustment for center was omitted when analyzing the effect of latitude.

### The type I interferon pathway is upregulated upon UVB phototherapy in MS patients

As it is unclear which molecular pathways mediate the beneficial effects of UVR in MS, PBMC samples of five UVB-treated MS patients from our 2014 pilot study were FACsorted for CD4 T-cells, CD8 T-cells, monocytes and B-cells, and subjected to RNAseq analysis (Fig. 5 A). For CD4 T-cells there were 268 genes that were regulated with *P*<0.05; 524 genes for CD8 T-cells, 411 genes for monocytes, and 407 genes for B-cells. Among these were previously described markers of vitD signaling, e. g. *VDR* (CD4 T-cells), *NR4A2, NR4A3* (CD4-, CD8 T-cells and monocytes) and *CD14* (monocytes) (Fig. 5 B-E). Furthermore, a first visual inspection suggested an enrichment of genes belonging to the type I interferon-family, including *IFITM1, IFITM2, IFITM3, MX1, IRF8, IRF7, IFI44L, IFIT2* and *IFIT3*, and this was most apparent in monocytes (Fig. 5 B-E). Regulation of exemplary genes for both pathways are plotted in Figure 5 (Fig. 5 F-I). A downregulation of the Aryl-hydrocarbon receptor-associated genes *AHR* and *TIPARP* was observed, as well. Next, to check whether the vitD- and the type I interferon-pathway were significantly enriched, permutation tests using reference gene-sets from *wikipathways* were performed. A significant enrichment of vitD-associated genes was found in CD8 T-cells (gene ratio (GR)= 0.108, *P*=0.002), monocytes (GR=0.079, *P*=0.018), and B-cells (GR=0.114, *P*=1×10^−4^). Furthermore, type I interferon-pathway-associated genes were significantly enriched in CD8 T-cells (GR=0.101, *P*=0.018), monocytes (GR=0.156, *P*=1×10^−6^), and B-cells (GR=0.083, *P*=0.027) (Fig. 5 J-M).

**Figure 5:**
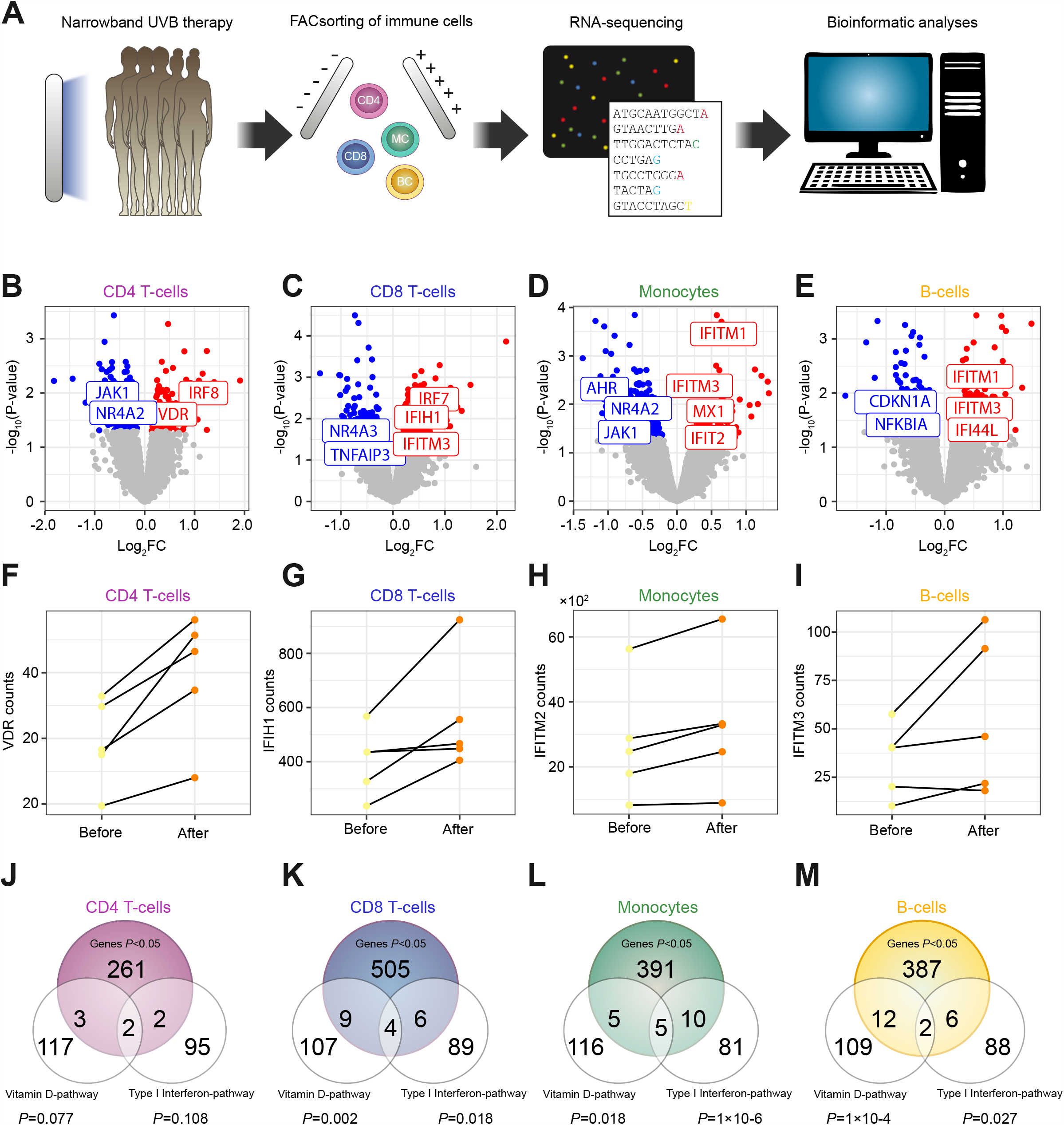
Transcriptional effects of UVB-phototherapy in immune cells. **A** Multiple sclerosis patients had been treated with UVB-phototherapy during the course of our 2014 pilot study (Breuer *et al*., 2014). PBMC Samples of five patients before and four weeks after phototherapy (10 samples in total) have been used to isolate CD4 T-cells that were subjected to RNAisolation and RNA-sequencing with subsequent bioinformatics analyses including differential expression analysis and gene set enrichment analysis (over-representation analysis). **B-E** Labeled, Volcano plots for CD4-, CD8 T-cells, monocytes and B-cells. **F-I** Before after plots for exemplary genes associated with either the vitD or the type I interferon pathway, regulated by photherapy in respective cell-types. **J-M** Venn diagrams for the overlap between significantly regulated genes in the respective cell types and the vitamin D and type I interferon gene-sets (extracted from wikipathways). *P*-values were calculated from distribution-free permutation-tests. The numbers for the reference gene sets refer to the number of genes belonging to the reference gene set and that have detectable expression in the respective cell type.

## Discussion

Low exposure to UVR and low vitD levels have been associated with the *development* of MS (5). It is still a topic of debate though, whether UVR, vitD and/or other UVR-dependent pathways, have an influence on *disease severity* and -*worsening over time*, as well. Most work has focused on vitD and only few studies investigated alternative UVR-mediated pathways. In the current study, data of two large, independent multicenter cohorts was used to test the association of latitude and vitD with MS disease severity. In this regard, modification of UVR-dependent effects by medication and sun-sensitivity-associated *MC1R* missense alleles was investigated, as well. To explore by which mechanisms UVR modulates MS severity next generation sequencing was performed to assess the UVB phototherapy-induced changes in immune cells in a small group of patients.

In line with previous studies, we observed lower disease severity in patients with higher vitD levels in the *NationMS* cohort (5). Expectedly, vitD levels were also strongly associated with latitude. Therefore, latitude itself is a sound proxy for sun exposure / vitD on a population level, especially if the sampling month is considered, as well. Consistently, latitude also showed a significant association with severity in the *NationMS* cohort. This means it is unlikely that reverse causation drives the association between vitD and disease severity. If there was reverse causation, the association between low vitD and increased severity would be expected, but there should be no association of high latitude (and therefore, low sun exposure) with increased severity. Although the absence of reverse causation does not imply a causative role for vitD since any UVR-induced metabolite that behaves similar to vitD would show the same pattern, this does support relevant effects of UVR in MS. In the *BIONAT* cohort vitD was also associated with reduced disability. Interestingly, the effect of latitude was only observed in patients who were not treated with IFN-β before. Since interactions between IFN-β treatment and vitD have been shown before (22, 23), it is likely that the normal association between latitude and vitD is altered by IFN-β treatment, which is also in line with the observation that patients with IFN-β therapy had significantly higher vitD levels compared to treatment-naïve patients in this study.

To uncover the UVR-induced transcriptomic changes in immune cells in MS patients, the effect of UVB phototherapy was assessed using next generation sequencing. Due to the limited sample size and the high false discovery rate burden, genes with *P*<0.05 were regarded as regulated and provided sensible results. The regulation of genes such as *NR4A2, NR4A3, CD14* and the gene for the vitamin D receptor (*VDR*), which have been reported multiple times to indicate a vitD response *(43, 44)* is in line with the profound vitD increase upon phototherapy that was previously demonstrated in these patients (12). As vitD counter-acts parathyroid hormone (PTH) expression (45), it is likely that the downregulation of the *NR4A*-family is due to a reduction of PTH, as PTH has been shown to increase *NR4A*-expression (46). This is also in line with the observed general enrichment of vitD-associated genes in permutation tests. Moreover, we identified regulation of the type I interferon pathway by phototherapy. As an upregulation of type I interferon-associated genes was observed in all donors of whom only two received IFN-β therapy, this relationship cannot be explained by IFN-β therapy, but instead is likely to be a direct result of phototherapy. Type I interferon induction was most apparent in monocytes – the natural producers of type I interferons in the context of pathogen control (47). A UVR dependent induction of type I interferons could also explain why the effect of latitude on disability in the *BIONAT* cohort was absent in IFN-β treated patients. If part of the effect of UVR is mediated directly through type I interferons, it is possible that IFN-β treatment could mask the effects of UVR / latitude. In fact, the type I interferon genes found to be regulated have previously been shown to be regulated upon IFN-β therapy in RRMS patients who were classified as IFN-β therapy responders (48, 49). The downregulation of *AHR* and *TIPARP*, known counter-actors of type I interferons-signaling, further supports a direct upregulation of type I interferons through phototherapy (50). Mechanistically, it is possible that the type I interferon-expression is modulated by vitD, as interactions between vitD and IFN-α/β have been described (22, 51). This is also in line with reports showing that vitD and IFN-β triggered gene expression partly overlaps (52).

Moreover, *in vitro-* and animal studies have proposed that UVR could also induce type I interferons via nucleic acid-damage and induction of damage associated molecular patterns (DAMPs) that can be sensed by toll-like receptors and the stimulator of interferon genes (STING) (53, 54). Interestingly, type I interferons were also shown to be induced in the skin of healthy human individuals upon UVR exposure, which lines up with the results from this study (55). If the type I interferon-pathway was indeed regulated by UVR, this could provide a link between MS and Lupus. In direct contrast to MS, in Lupus, an autoimmune disease that can manifest locally (e. g. cutaneous lupus erythematosus or CLE) or systemically (systemic lupus erythematosus or SLE), exposure to UVR is known to be disease-worsening (56). Furthermore, Lupus is known to be partly driven by type I interferons and patients often show an increased type I interferon blood-signature (57). Blockade of the type I interferon-pathway has been demonstrated to reduce disease severity in SLE (58). On the contrary, MS is associated with a reduced type I interferon signature and patients respond positively to IFN-β therapy (59).

Furthermore, although sun exposure seems to be beneficial for MS patients, it is also important to consider individual patient characteristics, such as skin type and sun sensitivity, and we therefore also investigated modulatory effects of sun-sensitivity-associated *MC1R* variants. In line with the concept of weak and strong *MC1R* variant alleles (24), only the high-penetrance variant rs1805008(C>T) showed a significant effect on self-reported skin reaction to sun exposure, whereas no association was found for rs885479(G>A) and rs2228479(G>A). The inversed effect of latitude on the number of Gd-enhancing lesions in carriers and non-carriers of the rs1805008 variant is in line with the observation of increased sun-sensitivity and a higher grade of inflammation induced through sun-exposure, combined with the reduced responsiveness to anti-inflammatory stimuli of α-melanocyte stimulating hormone in carriers of *MC1R* variant alleles (60, 61). The fact that this inversed association was not present across read-outs could be due to the fact that there was no evidence for correlation between the MSSS and the number of Gd-enhancing lesions. Furthermore, as the effects of *MC1R* in this study were only nominally significant, the results must be interpreted with care and require confirmation in larger cohorts.

In summary, this study provides evidence for an effect of UVR on MS severity. This argues for a recommendation of moderate sun-exposure for MS patients. Mechanistically, vitD is supported as one of the main mediators of UVR-effects, but a role for the type I interferon- and the MC1R-pathway is brought up, as well. Differential effects of UVR in carriers and non-carriers of *MC1R* variants were observed, suggesting detrimental effects of UVR in sun-sensitive patients. Therapy with IFN-β reduced the effect of latitude on severity that was observed in naïve patients, and the type I interferon pathway was upregulated by phototherapy in immune cells of MS patients. A direct upregulation of type I interferons through UVR would explain why no effect of latitude was observed in IFN-β treated patients. Therefore, this study thereby suggests type I interferon pathway as a novel and direct mediator of UVR-effects in MS. This also provides a framework to better understand the opposite effects of sun exposure in MS and the type I interferon-mediated and sun-sensitivity-associated disease Lupus.

## Data Availability

Data and R code is available from the authors upon reasonable request. Full results from RNAseq can be found in the supplementary material.

## Acknowledgements

The authors thank Nicole Hessler and Prof. Inke R. König (University of Lübeck, Germany) for their statistical contributions, and Prof. Markus Herrmann (University of Graz, Austria), Dr. Irene Pusceddu and Manuela Lucchiari (Bolzano Hospital, Italy) for 25(OH)D_3_ measurements; Lise Scandella for technical assistance for Toulouse; Petra Kotte, Barbara Meyring and Jila Abkari for technical assistance for Münster; and the genomics core facility of the medical faculty of Münster university for RNA sequencing.

## Authors’ contributions

**PO** conducted analyses, prepared figures, tables, and wrote the manuscript draft. **DG** contributed to statistical analyses and reviewed the chosen approaches. **JB** and **TSH** contributed to the analyses and literature research. **NM** and **KL** provided expertise in *MC1R* and skin biology and contributed to literature research. **BMM, TFMA, BH** and **TD** prepared and provided genetic data (*MC1R* SNPs) and expertise concerning population structure and genetic analysis. **GA** prepared patient data on behalf of the KKNMS and contributed to bioinformatical system administration. **RH, TK, LK, FZ, SGM** contributed to the clinical analysis approach and gave critical intellectual input. **NS** and **HW** designed the study, contributed to the analyses, literature research, provided funding, and wrote the manuscript. **RG** and **AS** were responsible for the study protocol, design and ethics implementation of the NationMS cohort study, and for measurement of serum vitD of NationMS samples. **CCG** and **ASM** analyzed data and provided critical intellectual input. **CL** coordinated MRI assessments across KKNMS centers. **SG, GGE, CL** and **MM** analyzed MRI data. **DB, BP, FB, IG** contributed and analyzed data from the BIONAT cohort. **AB, FTB, SB, HPH, MAF, RAL, FL, KLH, FP, MS, BT, HT, CW, FW, BW, UKZ** and **UZ** provided administrative, technical and material support. All authors contributed to the writing of the manuscript and provided critical revision to the text.

## Supplementary Information

Content:

- KKNMS study group members
- BIONAT study group members
- See .xlsx file for RNAseq supplementary information
- Supplementary Figure 1

## KKNMS study group members

Department of Neurology. Klinikum Augsburg. Augsburg. Germany

- Antonios Bayas
- Susanne Rothacher
- Stephanie Starke

Department of Neurology. Charité - Universitätsmedizin Berlin. Corporate member of Freie Universität Berlin. Humboldt-Universität zu Berlin and Berlin Institute of Health. Berlin. Germany

- Friedemann Paul
- Judith Bellmann-Strobl
- Janina Behrens
- Jan-Markus Dörr
- Rene Gieß
- Joseph Kuchling
- Ludwig Rasche

Department of Neurology. St. Josef-Hospital. Ruhr-University Bochum. Bochum. Germany

- Ralf Gold
- Andrew Chan
- Gisa Ellrichmann
- Anna Lena Fisse
- Anna Gahlen
- Thomas Grüter
- Aiden Haghikia
- Robert Hoepner
- Ümmügülsün Koc
- Carsten Lukas
- Jeremias Motte
- Kalliopi Pitarokoili
- Anke Salmen
- Ruth Schneider
- Joanna Schöllhammer
- Christoph Schroeder
- Björn Ambrosius
- Seray Demir

Department of Neurology. Medical Faculty. University of Düsseldorf. Düsseldorf. Germany

- Clemens Warnke
- Thomas Dehmel
- Kathleen Ingenhoven

Department of Neurology. University Hospital Erlangen. Friedrich-Alexander-University Erlangen-Nürnberg (FAU). Schwabachanlage 6. 91054. Erlangen. Germany

- Ralf Linker
- De-Hyung Lee
- Alexandra Lämmer
- Eva Sauer

MS Day Hospital and Institute of Neuroimmunology and Multiple Sclerosis. University Medical Center Hamburg-Eppendorf. Hamburg. Germany

- Christoph Heesen
- Jan-Patrick Stellmann

Clinical Neuroimmunology and Neurochemistry. Department of Neurology. Hannover Medical School and Centre for Systems Neuroscience. Hannover. Germany

- Martin Stangel
- Lena Boenig
- Stefan Gingele
- Martin Hümmert
- Philipp Schwenkenbecher
- Thomas Skripuletz
- Wolfram Suehs

Molecular Neuroimmunology Group Department of Neurology University of Heidelberg Heidelberg Germany

- Brigitte Wildemann
- Mirjam Korporal-Kuhnke
- Hanna Oßwald
- Alexander Schwarz
- Andrea Viehöver

Clinic for Neurology & Palliative Medicine. Cologne-Merheim. Cologne. Germany

- Volker Limmroth
- Kathrin Gerbershagen

Department of Neurology. University of Leipzig. Leipzig. Germany

- Florian Then Bergh
- Barbara Ettrich
- Steffi Gray
- Sarah Haars.
- Johannes Orthgieß
- Nicole Schwanitz
- Muriel Stoppe
- Astrid Unterlauft

Department of Neurology. University Medical Center of the Johannes Gutenberg University Mainz. Mainz. Germany

- Sandra Paryjas
- Stefan Bittner
- Vinzenz Fleischer
- Sergiu Groppa
- Felix Lüssi
- Johannes Piepgras
- Timo Uphaus

Department of Neurology. Philipps-University of Marburg. Marburg. Germany

- Björn Tackenberg
- Michael Pütz
- Christian Eienbröker
- Maria Seipelt

Institute of Clinical Neuroimmunology. Biomedical Center and University Hospitals. Ludwig-Maximilians-Universität München. Munich. Germany

- Reinhard Hohlfeld
- Tania Kümpfel
- Joachim Havla
- Ingrid Meinl
- Hannah Pellkofer
- Elisabeth Schuh

Department of Neurology. Klinikum rechts der Isar. Technische Universität München. Munich. Germany

- Bernhard Hemmer
- Lilian Aly
- Achim Berthele
- Viola Pongratz
- Kirsten Brinkhoff
- Dorothea Buck
- Christiane Gasperi
- Mirjam Hermisson
- Muna-Miriam Hoshi
- Miriam Kaminski
- Ana Klein
- Benjamin Knier
- Markus Kowarik
- Helena Kronsbein
- Klaus Lehmann Horn
- Meike Mitsdörffer
- Verena Pernpeintner
- Veit Rothhammer
- Andrea Schweikert
- Rebecca Selter

Department of Neuroradiology, Klinikum rechts der Isar, Medical Faculty, Technical University of Munich, Germany

- Mark Mühlau
- Claus Zimmer
- Jan Kirschke

Max Planck Institute of Psychiatry. Munich. Germany; Neurological Clinic Cham. Sana Kliniken des Landkreises Cham GmbH Park. Germany

- Frank Weber
- Heike Staufer
- Matthias Knop
- Sandra Nischwitz
- Philipp Sämann

Department of Neurology with Institute of Translational Neurology. University of Münster. Münster. Germany

- Heinz Wiendl
- Sven Meuth
- Luisa Klotz
- Gerd Meyer zu Hörste
- Julia Krämer
- Lena Schünemann
- Catharina Gross
- Steffen Pfeuffer
- Tobias Ruck
- Selma Belgriri
- Alexander Buchheister
- Nora Bünger
- Kerstin Göbel
- Lucienne Kirstein
- Nico Melzer
- Ole Simon
- Antje Echterhoff

Department of Neurology. Neuroimmunological Section. University of Rostock. Rostock. Germany

- Uwe Zettl
- Alexander Winkelmann

Institute of Medical Psychology and Behavioral Neurobiology. University of Tübingen. Germany; International Max Planck Research School (IMPRS) for Cognitive and Systems Neuroscience. Tübingen. Germany

- Ulf Ziemann
- Ahmed Abdelhak
- Markus Kowarik
- Markus Krumbholz
- Margarete Paech
- Christoph Ruschil
- Maria-Ioanna Stefanou.
- Johannes Tünnerhoff
- Lena Zeltner
- Hajera Sheikh

Department of Neurology. University of Ulm. Ulm. Germany.

- Hayrettin Tumani
- Tanja Fangerau
- Florian Lauda
- Daniela Rau
- Daniela Taranu
- André Huss

## BIONAT study group members

- David Brassat. MD. PhD - CRC SEP – Neurosciences – CHU Toulouse and CPTP INSERM U1043 CNRS U5282 – Université Toulouse III – France
- Béatrice Pignolet. PhD - CRC SEP – Neurosciences – CHU Toulouse and CPTP INSERM U1043 CNRS U5282 –France
- Florence Bucciarelli. MSc - CRC SEP – Neurosciences – CHU Toulouse and CPTP INSERM U1043 CNRS U5282. France
- Lise Scandella - CRC SEP – Neurosciences – CHU Toulouse and CPTP INSERM U1043 CNRS U5282 – Université Toulouse III – France
- Christine Lebrun-Frenay. MD - CRCSEP Côte d’Azur. CHU de Nice Pasteur. Université Nice Côte d’Azur. Nice. France (sample and clinical data collection)
- Marc Debouverie. MD. PhD - Department of Neurology. Nancy University Hospital. F-54035 Nancy. France
- Sophie Pittion-Vouyovitch. MD – Department of Neurology. Nancy University Hospital. F-54035 Nancy. France (sample and clinical data collection)
- Bruno Brochet - CHU Pellegrin Bordeaux. France (sample and clinical data collection)
- Aurelie Ruet - CHU Pellegrin Bordeaux. France (sample and clinical data collection)
- Gilles Defer. MD. PhD - CHU Caen - France (sample and clinical data collection)
- Nathalie Derache - CHU Caen – France (sample and clinical data collection)
- Jérôme de Sèze. MD. PhD - CHU Strasbourg. France (sample and clinical data collection)
- David Laplaud. MD. PhD – CHU Nantes. France (sample and clinical data collection)
- Sandrine Wiertlewski. MD - CHU Nantes. France (sample and clinical data collection)
- Oliver Casez. MD – CHU Grenoble. France (sample and clinical data collection)
- Pierre Clavelou. MD. PhD - Neurology. CHU Montpied. Clermont-Ferrand. France
- Pierre Labauge. MD. PhD – CHU Montpellier. France (sample and clinical data collection)
- Jean Pelletier. MD. PhD - APHM. Hôpital de la Timone. Pôle de Neurosciences Cliniques. Service de neurologie. CRCSEP Marseille. France
- Audrey Rico. MD - APHM. Hôpital de la Timone. Pôle de Neurosciences Cliniques. Service de neurologie. CRCSEP Marseille. France (sample and clinical data collection)
- Sandra Vukusic. MD. PhD - CHU Lyon Bron. France (sample and clinical data collection)
- Olivier Outteryck. MD (Neurology. Université de Lille. France; sample collection and clinical data collection).
- Jean-Claude Ongagna. MD (Neurology. Hôpital Civil. Strasbourg. France; sample collection and clinical data collection).
- Jean-Christophe Ouallet. MD (Neurology. CHU Pellegrin. Bordeaux. France; sample collection and clinical data collection).
- Prof Patrick Hautecoeur. MD (Neurology. CH St Vincent. GHICL. Lille. France; sample collection and clinical data collection).
- Prof Ayman Tourbah. MD (Neurology. CHU Reims. France; sample collection and clinical data collection).
- Giovanni Castelnovo. MD (Neurology. CHU Nîmes. France; sample collection and clinical data collection).
- Eric Berger. MD (Neurology. CHU Besançon. Besançon. France; sample collection and clinical data collection).
- Hélène Zéphir. MD (Neurology. Université de Lille. France; sample collection and clinical data collection).
- Philippe Cabre. MD (Neurology. CHU Fort de France. Fort de France. France; sample collection and clinical data collection).
- Prof William Camu. MD (Neurology. CHU Montpellier. France; sample collection and clinical data collection).
- Prof Eric Thouvenot. MD (Neurology. CHU Nîımes. Nîmes. France; sample collection and clinical data collection).
- Prof. Thibault Moreau. MD (Neurology. CHU Dijon. Dijon. France; sample collection and clinical data collection).
- Agnès Fromont. MD (Neurology. CHU Dijon. Dijon. France; sample collection and clinical data collection).
- Caroline Papeix. MD (Neurology. Hôpital de la Salpétrière. Paris. France; sample collection and clinical data collection.
- Prof Catherine Lubetzki. MD (Neurology. Hôpital de la Salpétrière. Paris. France; sample collection and clinical data collection).
- Prof Patrick Vermersch. MD (Neurology. Université de Lille. France; sample collection and clinical data collection).
- Mikael Cohen. MD (Neurology. Hôpital Pasteur. Nice. France; sample collection and clinical data collection).
- Prof. Lucien Rumbach. MD (Neurology. CHU Besançon. Besançon. France; sample collection and clinical data collection).

**Supplementary Figure S1:**
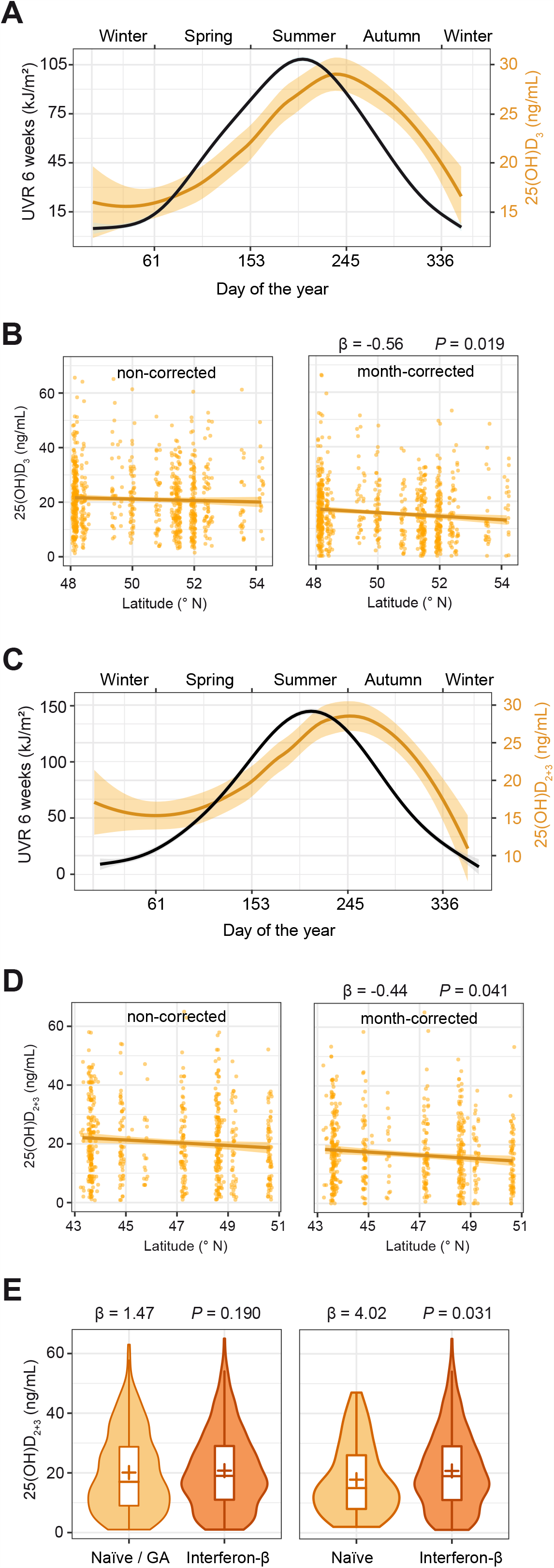
Effect of sampling month, latitude and medication on serum vitamin D. **A** Influence of recent UVR exposure and sampling month on serum vitamin D levels in the *NationMS* cohort. Black loess curve depicts recent UVR exposure at a patients’ medical center (6 weeks before sampling; sum of daily UVR intensities as extracted from NASA’s OMI dataset on erythemal UVR at a 1×1° resolution in latitude and longitude) and corresponds to the left y-axis. Orange loess curve depicts serum vitamin D levels and corresponds to the right y-axis. **B** Dot-plot showing the influence of latitude on serum vitamin D with a least-squares regression line ± standard error. Values in the right panel were corrected for sampling month by first regressing sampling month on serum vitamin D and using the model residuals plus the model intercept as month-corrected vitamin D levels for the *NationMS* cohort. **C** Influence of recent UVR exposure and sampling month on serum vitamin D levels in the *BIONAT* cohort. Black loess curve depicts recent UVR exposure at a patients’ medical and corresponds to the left y-axis. Orange loess curve depicts serum vitamin D levels and corresponds to the right y-axis. **D** Dot-plot showing the influence of latitude on serum vitamin D with a least-squares regression line ± standard error. Values in the right panel were corrected for sampling month by first regressing sampling month on serum vitamin D and using the model residuals plus the model intercept as month-corrected vitamin D levels for the *BIONAT* cohort. **E** Violin-plot showing serum vitamin D-levels with regard to prior medication for the *BIONAT* cohort.

